# COPD with elevated sputum group 2 innate lymphoid cells is characterized by severe disease

**DOI:** 10.1101/2023.11.21.23298837

**Authors:** Cameron H. Flayer, Angela L. Linderholm, Moyar Q. Ge, Maya Juarez, Lisa Franzi, Tina Tham, Melissa Teuber, Shu-Yi Liao, Michael Schivo, Brooks Kuhn, Amir Zeki, Angela Haczku

## Abstract

**Rationale:** Pulmonary innate immune cells play a central role in the initiation and perpetuation of chronic obstructive pulmonary disease (COPD), however the precise mechanisms that orchestrate the development and severity of COPD are poorly understood.

**Objectives:** We hypothesized that the recently described family of innate lymphoid cells (ILCs) play an important role in COPD.

**Methods:** Subjects with COPD and healthy controls were clinically evaluated, and their sputum samples were assessed by flow cytometry. A mouse model of spontaneous COPD [genetically deficient in surfactant protein-D (SP-D^-/-^)] and ozone (O_3_) exposure were used to examine the mechanism by which lack of functional SP-D may skew ILC2s to produce IL-17A in combination with IL-5 and IL-13, leading to a mixed inflammatory profile and more severe disease.

**Measurements and Main Results:** COPD was characterized by poor spirometry, sputum inflammation, and the emergence of sputum GATA3^+^ ILCs (ILC2s), but not T-bet^+^ ILCs (ILC1s) nor RORγt^+^ ILCs (ILC3s). COPD subjects with elevated sputum ILC2s (the ILC2^high^ group) had worse spirometry and sputum neutrophilia and eosinophilia than healthy and ILC2^low^ subjects. This was associated with the presence of dual-positive IL-5^+^IL-17A^+^ and IL-13^+^IL-17A^+^ ILCs and nonfunctional SP-D in the sputum in ILC2^high^ subjects. SP-D^-/-^ mice showed spontaneous airway neutrophilia. Lack of SP-D in the mouse lung licensed ILC2s to produce IL-17A, which was dose-dependently inhibited by recombinant SP-D. SP-D^-/-^ mice showed enhanced susceptibility to O_3_-induced airway neutrophilia, which was associated with the emergence of inflammatory IL-13^+^IL-17A^+^ ILCs.

**Conclusions:** We report that the presence of sputum ILC2s predicts the severity of COPD, and unravel a novel pathway of IL-17A plasticity in lung ILC2s, prevented by the immunomodulatory protein SP-D.

## INTRODUCTION

While chronic obstructive pulmonary disease (COPD) continues to be a leading cause of mortality worldwide (1), the mechanisms of innate immunity central to disease pathogenesis remain uncertain. A previously overlooked family of cells, the recently described innate lymphoid cells (ILCs), have emerged as a pivotal innate immune population in a variety of pulmonary diseases (2, 3). This family can be broadly classified as group 1 ILCs (ILC1s) that are labeled by T-bet expression and IFNγ, group 2 ILC2s that are defined by GATA3 expression and IL-5/IL-13, and group 3 ILC3s that are characterized by RORγt expression and IL-17A (2, 4). In asthma, group 2 innate lymphoid cells that produce IL-5 and IL-13 were singled out for their propensity to initiate airway inflammation and impair lung function (5, 6). In COPD however, it was found that ILC2s gain an ILC1-like phenotype, acquiring T-bet expression and producing IFNγ (7). In patients with COPD, the frequency of peripheral blood ILC1s increased and correlated to disease severity, while the frequency of peripheral blood ILC2s decreased (7). In a study of human lung tissue, COPD subjects showed an elevated frequency of ILC3s, not ILC1s nor ILC2s, compared to healthy controls (8). These conflicting data derived from different tissue compartments suggest a role for ILCs in COPD, though the relevance of specific ILC subsets to disease severity and the underlying mechanisms of their activation remain uncertain. How might ILCs get highly activated and contribute to disease pathogenesis in COPD?

The lung collectin surfactant protein-D (SP-D) is an immunoprotective protein important to COPD pathogenesis (9–15). Indeed, airway SP-D is diminished in subjects with COPD, both in former and current smokers (16). A recent study found in smokers, serum SP-D levels increased with lung function decline (17). Mice genetically deficient in SP-D (SP-D^-/-^) develop histologically and physiologically quantifiable emphysema, a hallmark of COPD (18). While these mice additionally develop spontaneous airway inflammation and activation of pulmonary innate immune cells, the relationship between SP-D and ILCs has not been characterized. Given these observations, here we sought to clarify the role of ILCs in COPD disease severity and understand how SP-D impacts disease pathogenesis and ILC activation.

To address our questions, we recruited subjects with COPD and healthy controls from the University of California, Davis pulmonary and primary care clinics. We explored the relationship between sputum ILCs and clinical parameters including spirometry, COPD assessment test questionnaire, sputum inflammation, and SP-D levels. With these observations, we defined the mechanisms underlying the relationship between SP-D and ILC2s in SP-D^-/-^ mice and the impact of SP-D on ILC2s *ex vivo*.

## METHODS

### Human subjects

Patients with COPD (n=15) and non-smoking healthy volunteers (n=12) were recruited from the UC Davis pulmonary and primary care clinics. Details on inclusion and exclusion criteria are provided in an online supplement.

Subjects that met eligibility criteria were consented and invited to enroll in the study. During a single study visit, the following procedures were conducted: review of medical record, physical examination, pulmonary function testing (PFT), health related quality of life surveys [modified medical research council scale (mMRC) and COPD assessment test (CAT)], venipuncture for collection of 20 mL blood, sputum induction by inhalation of 3% hypertonic saline, and 6-minute walk test. Healthy volunteers underwent all procedures except for the physical examination. For COPD patients, a physical examination, PFT, and 6-minute walk test performed within 2 months of the initial study visit was acceptable. This study was reviewed and approved by the University of California, Davis Institutional Review Board.

### Sputum

10% Sputolysin reagent (MilliporeSigma, Burlington, MA) was added 1:1 weight/volume to induced sputum specimens. The sample was then placed in a 37°C shaking water bath for 15 minutes. Every 5 minutes, the sample was removed to be pipette mixed. Undiluted Sputolysin reagent was added as necessary if sample homogenization was poor. Sputum cells were strained through a 40 µm Olympus Advanced cell strainer (Genesee Scientific, San Diego, CA) before cell count by the Countess™ Automated Cell Counter. Approximately 32x10^3^ cells were centrifuged at 500 rpm for 5 minutes onto a microscope slide, then stained via the Shandon Kwik-Diff staining kit (Thermo Fisher Scientific, Waltham, MA). 2 cytospins were made for each subject. The remaining cells were apportioned for flow cytometry, while the sputum supernatant was aliquoted and snap frozen for storage at -80°C.

Cytospins were evaluated for the presence of macrophages/monocytes (M0), lymphocytes (LC), neutrophils (NP), eosinophils (EP), epithelial cells, and oral squamous cells. At minimum, 200 cells were counted per cytospin and differential cell percentages were averaged between cytospins for each subject. Sputum samples were deemed poor quality if >40% of the cytospin was squamous cells. This quality control criterium required n=4 healthy and n=5 COPD subjects to be excluded from the final analysis of sputum cells and supernatant, as well as ILC2^high^ and ILC2^low^ grouping.

### Mice

The experimental animals used in this study were housed under pathogen-free conditions with free access to food and water on a 12 hour light-dark cycle. C57BL/6 wild-type (WT), Balb/c, and Rag2/γc^-/-^ mice were obtained from The Jackson Laboratory (Sacramento, CA) and bred in house. SP-D^-/-^ mice on the C57BL/6 background were a gift from Drs. Samuel Hawgood, University of California, San Francisco, and Francis Poulain, University of California, Davis and bred in house. Experiments were conducted on 6-10 week old male mice. The University of California, Davis Institutional Animal Care and Use Committee approved the experimental protocol.

Mice were euthanized naïve or exposed to 3 ppm ozone (O3) or air for 2 hours, then euthanized 12 hours later. Bronchoalveolar lavage (BAL) fluid and lungs were harvested to measure airway neutrophilia and lung ILC2s, respectively. BAL was obtained by instilling 2.7 mL phosphate-buffered saline (PBS) into the lungs (in 3 instillations) via tracheal cannula to collect cells and proteins of interest. Lungs were excised and single cell suspensions were made by digestion in Liberase™ (Roche, Basel, Switzerland) for 40 minutes at 37° C. Cells were strained using a 70 µm cell strainer. Red blood cells were lysed, then strained a second time through a 70 µm strainer. BAL and lung cells were counted using the Countess™ Automated Cell Counter. Cell-free BAL supernatant was stored at -80°C for cytokine analysis.

For adoptive transfer studies, 250 µg αIL-17A or αIgG1 monoclonal antibodies were injected intraperitoneal (I.P.) into Rag2/γc^-/-^ mice at 0, 24, and 48 hours. ILC2s were expanded *ex vivo* from donor WT lungs in 30 ng/mL IL-2+IL-7+IL-33 for 2 weeks, then 10^5^ ILC2s were intravenously (I.V.) injected into the tail vein of recipient Rag2/γc^-/-^ mice at 32 hours. 24 hours later (56 hours after the first monoclonal antibody injection), mice were exposed to air or O_3_ and then euthanized 12 hours later. BAL was harvested to measure airway neutrophilia.

### Flow cytometry antibodies, staining, and analysis

For mouse studies, Fixable Viability Stain 510 (BD Biosciences, San Jose, CA) was used following manufacturer’s instructions. All anti-mouse antibodies were purchased from BioLegend (San Diego, CA), BD Biosciences (San Jose, CA), and eBioscience (San Diego, CA). A complete list of anti-mouse antibodies are provided in an online supplement.

For human studies, Zombie Green fixable viability kit (BioLegend, San Diego, CA) was used following manufacturer’s instructions. All anti-human antibodies were purchased from BioLegend (San Diego, CA), BD Biosciences (San Jose, CA), and Miltenyi Biotec (Auburn, CA). A complete list of anti-human antibodies are provided in an online supplement.

Flow cytometric data was collected on a LSRFortessa™ (BD Biosciences, San Jose, California) with FACS Diva™ software (BD Biosciences, San Jose, California) and analyzed using FlowJo software (FlowJo, LLC, Ashland, Oregon). Cells were sorted using a FACSAria II™ (BD Biosciences, San Jose, California).

For intracellular cytokine detection from mouse ILC2s in the lung, cells were restimulated in 12.5 ng/mL phorbol 12-myristate 13-acetate (PMA, MilliporeSigma Burlington, MA), 500 ng/mL Ionomycin (MilliporeSigma, Burlington, MA) and 3 µM Monensin (MilliporeSigma, Burlington, MA) for 6 hours at 37° C. Lung cell suspensions were incubated in surface antigens for 20 minutes at 4° C in the dark. Cells were fixed and permeabilized using the FoxP3 perm/fix kit (BD Biosciences, San Jose, CA), then incubated in intracellular antigens for 30 minutes at 4° C in the dark. FMO or Lineage^-^ Thy1.2^-^ cells were used as technical controls.

For intracellular cytokine and transcription factor detection from human ILCs in blood and sputum, freshly isolated sputum and PBMC cell suspensions were incubated in surface antigens for 30 minutes on a shaker at room temperature in the dark. Cells were fixed and permeabilized using the BD Cytofix/Cytoperm kit (BD Biosciences, San Jose, CA), then incubated in intracellular antigens for 30 minutes on a shaker at room temperature in the dark. FMO cells were used as technical controls.

### ILC2 culture (*ex vivo*)

Recombinant mouse IL-2, IL-7, and IL-33 were purchased from Peprotech (Rocky Hill, NJ).

ILC2 were isolated from the lungs of C57BL/6 and SP-D^-/-^ mice as CD45^+^Lineage^-^ Thy1.2^+^CD127^+^CD25^+^ST2^+^ cells (lineage markers – CD3, CD4 CD5, CD11c, GR-1, B220, DX5) and cultured in non-treated U-bottom 96 well-plates. ILC2s were plated at 2,000 cells/well and stimulated in “ILC2 complete medium” (RPMI (ThermoFisher Scientific, Waltham, MA), 10% fetal bovine serum (Genesee Scientific, San Diego, CA), 1% penicillin/streptomycin (Genesee Scientific, San Diego, CA) and 10 ng/mL IL-2, IL-7, and IL-33). For culture experiments with recombinant SP-D, ILC2s were plated at 5,000 cells/well and expanded for 21 days in ILC2 complete medium, then rested in RPMI-alone for 16 hours. Following a rest period, ILC2s were stimulated for 3 days at 2,000 cells/well with cytokines and 0, 2, 10, or 20 µg/mL SP-D. At the time of harvest, cell free supernatant was collected for cytokine analysis (ELISA). Mouse recombinant SP-D (mrSP-D) was purchased from Sino Biological (Beijing, China).

### ELISA and Luminex Assay

BAL KC was detected via Mouse CXCL1/KC DuoSet ELISA (R&D Systems Minneapolis, Minnesota) following the manufacturer’s protocol. IL-17A DuoSet ELISA (R&D Systems, Minneapolis, MN) was detected in cell-free ILC2 culture supernatant following the standard protocol. BAL cytokines were detected in naïve C57BL/6 and SP-D^-/-^ mice via Luminex assay (Roche, Little Falls, NJ). The standard assay protocols supplied by the manufacturer were followed.

Human sputum IL-8 and SP-D was determined using a Luminex® magnetic bead-based premixed multiplex assay (R&D Systems, Minneapolis, MN) according to the manufacturer’s protocol.

### Western Blot

Sputum SP-D was detected via Native-PAGE western blot. Total protein in the BAL was quantified via Pierce BCA Protein Assay Kit (Thermo Fisher Scientific, Waltham, MA). 3.125 µg protein was run under native gel electrophoresis conditions using NuPAGE™ 1x Tris-Glycine Native Running Buffer (Thermo Fisher Scientific, Waltham, MA) at 150V for 120 minutes. The gel was transferred to a mini Nitrocellulose membrane using the iBlot® gel transfer mini stacks (Thermo Fisher Scientific, Waltham, MA). The membrane was probed with mouse anti-SP-D 1:1000 (in-house 10F6E12 monoclonal) followed by goat anti-mouse conjugated HRP 1:3000 (Santa Cruz Biotechnology, Santa Cruz, CA), then activated via Pierce™ ECL Western Blotting Substrate (Thermo Fisher Scientific, Waltham, MA.

### qPCR

Total RNA was extracted from the whole lung with TRIzol Reagent (Thermo Fisher Scientific, Waltham, MA). cDNA was reverse-transcribed using the QuantiTect Reverse Transcription Kit (Qiagen, Hilden, Germany). qPCR was performed using Fast SYBR® Green Master Mix (Thermo Fisher Scientific, Waltham, MA) on a ViiA 7 Real-Time PCR System (Thermo Fisher Scientific, Waltham, MA). Fold change was calculated using the ΔΔC_t_ method, first normalizing values to *β*-*actin*, then to the treatment control. Primer sequences are included in the online supplement.

### Data Analysis

Statistical analyses are indicated in each figure legend. Student’s *t*-test, One-way ANOVA with Bonferroni’s multiple comparison’s test, Two-way ANOVA with Bonferroni’s multiple comparisons test, and Linear Regression were applied using Graphpad Prism v7 (Graphpad Software Inc, La Jolla, CA) throughout the study. We performed regression correction for age. *p<0.05, **p<0.01, ***p<0.001, #p<0.05 were used to determine significance. Asterisks were used to denote statistically significant differences between groups indicated in the figure legends (i.e. healthy vs. COPD, WT vs. SP-D^-/-^, and air vs. O_3)_. In O_3_ exposure experiments utilizing C57BL/6 and SP-D^-/-^ mice, hashmarks show interstrain differences (i.e. WT vs. SP-D^-/-^). The human study was conducted over a period of 4 months, with a maximum of 3 subjects enrolled and processed per day. In the mechanistic mouse studies, experiments were independently performed 2-3 times.

## RESULTS

### COPD sputum samples had significantly increased GATA3^+^ ILC2 counts

To examine the relationship between sputum ILCs and SP-D in COPD severity, we recruited COPD patients and healthy volunteers from the UC Davis pulmonary and primary care clinics. Subjects were clinically evaluated, and spirometry, sputum induction, and blood collection were performed. Pre-bronchodilator forced expiratory volume in 1 second (FEV_1_) and forced vital capacity (FVC), % of predicted values and ratio of FEV_1_ over FVC were collected from healthy and COPD subjects.

COPD subjects had decreased predicted percentages of FEV_1_, forced vital capacity (FVC), and FEV_1/_FVC (**Fig. 1A**). This corresponded to significantly elevated neutrophils and eosinophils in the sputum of patients with COPD compared to healthy controls (**Fig. 1B and 1C**). These clinical parameters are consistent with COPD, suggesting our cohort was representative of a broader population. To define the role ILCs in COPD severity, we phenotypically characterized and measured ILCs by flow cytometry of sputum cell suspensions. We identified ILCs as live lymphocytes (by forward and side scatter) that were Lineage^-^CD127^+^ cells and intracellular expression of IL-5, IL-13, IL-17A, and IFNγ. There were significantly more ILCs in COPD subjects compared to healthy subjects (**Fig. 1D**). The expression of transcription factors T-bet^+^ (ILC1s), GATA3^+^ (ILC2s), and RORγt^+^ (ILC3s) were also measured, surprisingly we observed an increase in the proportion of ILCs that expressed GATA3 in the sputum of subjects with COPD compared to healthy volunteers (**Fig. 1E and 1F**). Given these data, we speculated that GATA3^+^ ILCs (ILC2s) were relevant to the severity of COPD.

**Figure 1.**
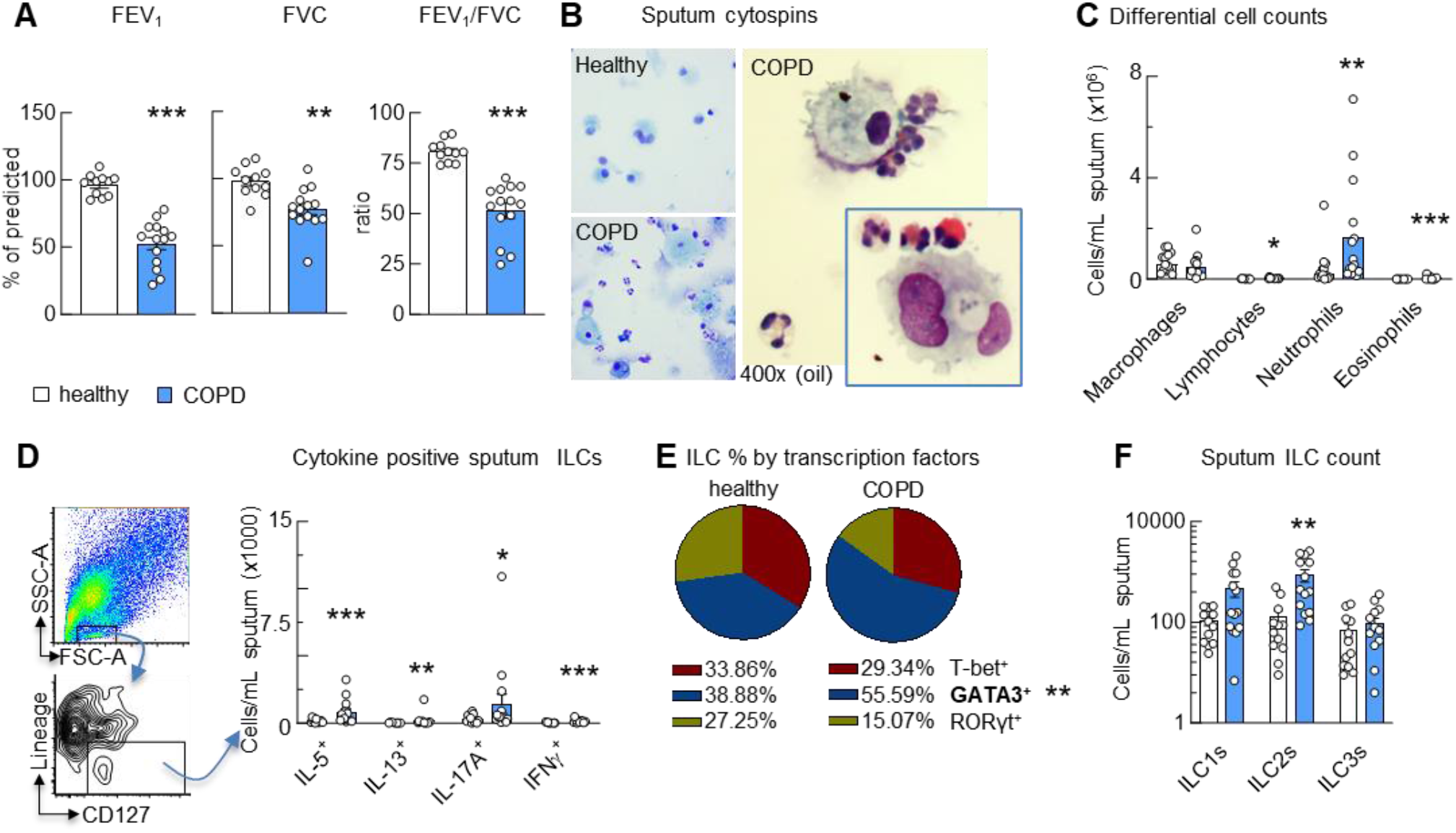
COPD sputum samples had significantly increased GATA3^+^ ILC2 counts. **(A)** Pre-bronchodilator forced expiratory volume in 1 second (FEV_1_) and forced vital capacity (FVC), % of predicted values and ratio of FEV_1_ over FVC of healthy and COPD subjects. **(B)** Cytospins of induced sputum cells from healthy (top) and COPD subjects were stained with Kwik-Diff. The enlarged images illustrate a macrophage surrounded by neutrophils; the insert shows a multi-nucleated macrophage, a neutrophil and two eosinophilic granulocytes. **(C)** Macrophages, lymphocytes, neutrophils, eosinophils and epithelial cells (not shown) were counted on at least 4 high-powered fields of 2 cytospin slides/subject. Percent differential counts were multiplied by the total sputum cell count/mL. **(D)** Identification of innate lymphoid cells [ILCs; live Lineage^-^CD127^+^ (lineage markers: CD3, CD14, CD16, CD19, CD20, CD56)] by flow cytometry in sputum and intracellular expression of IL-5, IL-13, IL-17A, and IFNγ (ILCs/mL sputum). Dotted line indicates background. **(E)** Pie chart average % values of ILCs that were T-bet^+^, GATA3^+^, and RORγt^+^. **p<0.01 (healthy (n=12) vs. COPD (n=15). **(F)** Counts of T-bet^+^ (ILC1s), GATA3^+^ (ILC2s), and RORγt^+^ (ILC3s), cells/mL sputum. **(A, C-D, F):** Mean±SEM of n=12-16 healthy, n=15-20 COPD. *p<0.05, **p<0.01, ***p<0.001, Mann-Whitney multiple comparisons test with FDR.

### COPD subjects with elevated sputum ILC2s (ILC2^high^) have increased numbers of inflammatory cells and IL-17+ ILC2s

We stratified COPD patients into “ILC2^high^” and “ILC2^low^” groups by the upper limit of the 95% confidence interval of sputum ILC2 count for healthy controls. Thus, COPD subjects with ILC2s >1427 cells/mL sputum were ILC2^high^ (n=8), while subjects with ILC2s <1427 cells/mL sputum were ILC2^low^ (n=7) (**Fig. 2A**). ILC2^high^ subjects additionally had significantly elevated sputum neutrophilia and increased eosinophilia compared to ILC2^low^ subjects (**Fig. 2B**). Correlations between ILC2 counts (dark blue circles) and eosinophils, neutrophils and FEV_1_ were made by linear regression using sputum samples from subjects with ILC2^high^ sputum (**Fig 2C**). These data suggest that the emergence of ILC2s in the sputum of COPD subjects is related to the severity of lung function decline, quality of life, and sputum inflammation. Interestingly, these observations also link sputum ILC2s to neutrophils and eosinophils. How could ILC2s be related to these inflammatory cells that are driven to the lung and maintained by different chemokines and cytokines?

**Figure 2.**
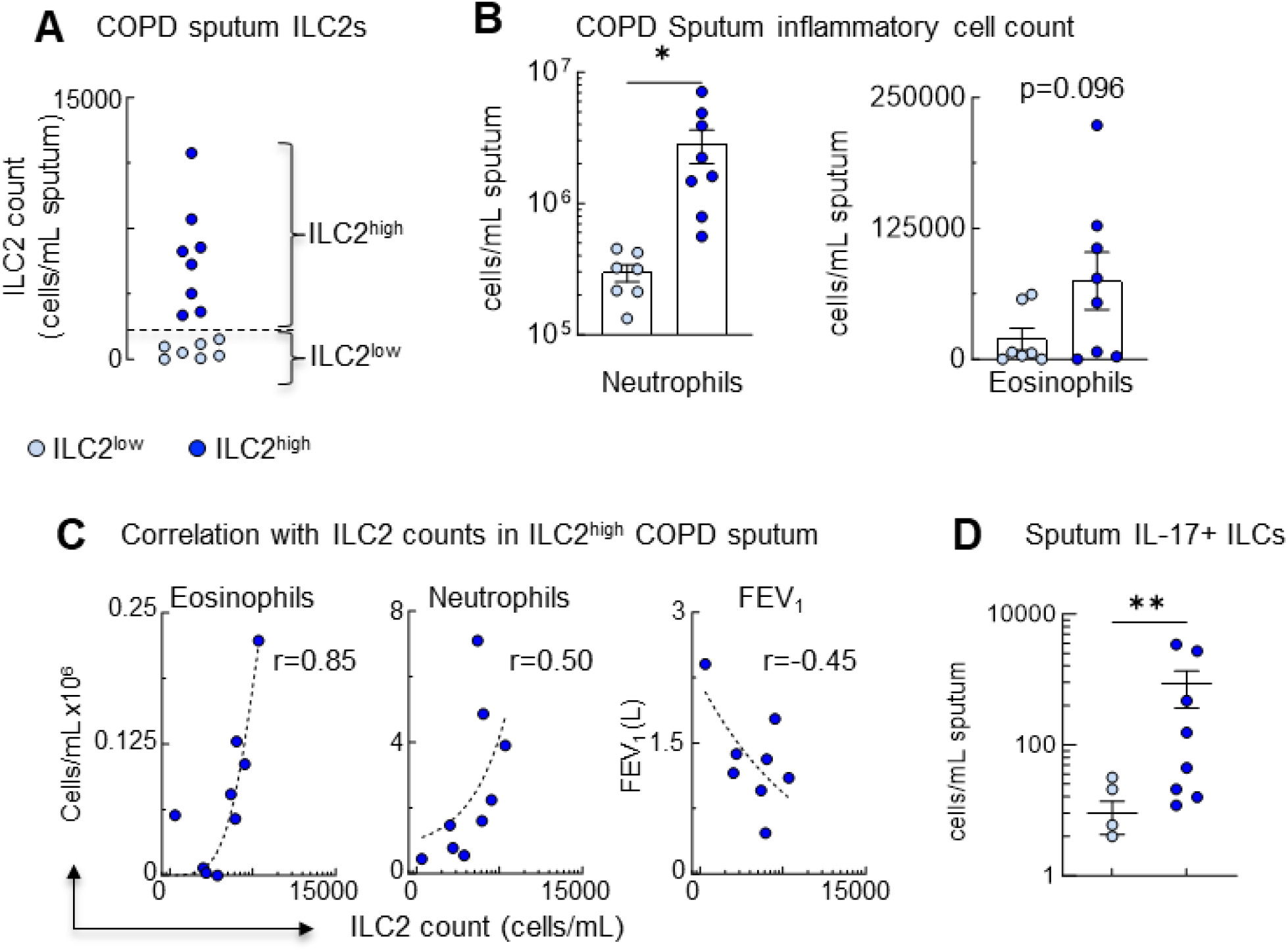
COPD subjects with elevated sputum ILC2s (ILC2^high^) had increased numbers of inflammatory cells and IL-17+ ILC2s. **(A)** COPD subjects were stratified according to their sputum ILC2 counts into ILC2^low^ (light blue circles) and ILC2^high^ (dark blue circles) groups, determined by the upper limit of the 95% confidence interval (1500 cells/mL sputum; dotted line). 9 of 15 COPD subjects had ILC2^high^ Sputum. **(B)** Sputum neutrophil and eosinophil counts of ILC2^low^ (light blue bar) and ILC2^high^ (dark blue bar) subjects (cells/mL sputum). **(C)** Correlations between ILC2 counts (dark blue circles) and eosinophils, neutrophils and FEV_1_ were made by linear regression using sputum samples from subjects with ILC2^high^ sputum (r: Pearson’s correlation coefficient). ILC2^low^ sputum sampes (light blue circles) were not included in the correlation. **(D)** ILC2s gated on GATA3^+^ expression from [ILCs; live Lineage^-^CD127^+^ (lineage markers: CD3, CD14, CD16, CD19, CD20, CD56)] were assessed for intracellular IL-17. **(A-B, D)** Mean±SEM of n=15-20 COPD patients. **(B, D)** Mean±SEM of ILC2^low^ and ILC2^high^ samples from n=7 and n=8 COPD subjects, respectively; * p<0.05, ** p<0.01, Mann-Whitney multiple comparisons test with FDR.

IL-5 and IL-13 are key ILC2 cytokines, and IL-5 is particularly relevant given its ability to prevent eosinophil apoptosis (2). On the other hand, RORγt ILC3s canonically produce IL-17A which plays a key role in initiating IL-8 production by epithelial cells to induce neutrophilia (2). There is evidence from prior studies that ILC2s can co-express IL-13 and IL-17A under specific inflammatory conditions, and these cells were termed inflammatory ILC2s (19, 20). Thus, to understand if ILC2s could potentially regulate both neutrophilia and eosinophilia by co-expressing IL-5 or IL-13 with IL-17A, we quantified ILC2s gated on GATA3^+^ expression for intracellular IL17^+^ in the sputum and found that ILC2^high^ subjects displayed elevated GATA3^+^IL-17A^+^ ILCs (**Fig. 2D**).

### Characteristics of healthy subjects and patients with ILC2^low^ and ILC2^high^ COPD sputum

The characteristics of healthy subjects (n=12) and patients with COPD, separated by ILC2^low^ (n=7) and ILC2^high^ (n=8) classifications, details of stratification are described above, are displayed in **Table 1**. Subjects with COPD, regardless of ILC2 status were significantly older and had a history of smoking, decreased specific oxygen (SpO_2_), increased modified medical research council scale (mMRC) and COPD assessment test (CAT) scores, and covered fewer meters in a 6-minute walk test compared to healthy volunteers (**Table 1**). COPD ILC2^high^ patients had higher CAT scores compared to COPD ILC2^low^ patients but were otherwise not significantly different. Neutrophils were also significantly increased in the peripheral blood of COPD ILC2^low^ and ILC2^high^ patients compared to healthy controls (**Table 1**). We recognized that our COPD cohort was significantly older than the healthy controls (**Table 1**).

**Table 1.**
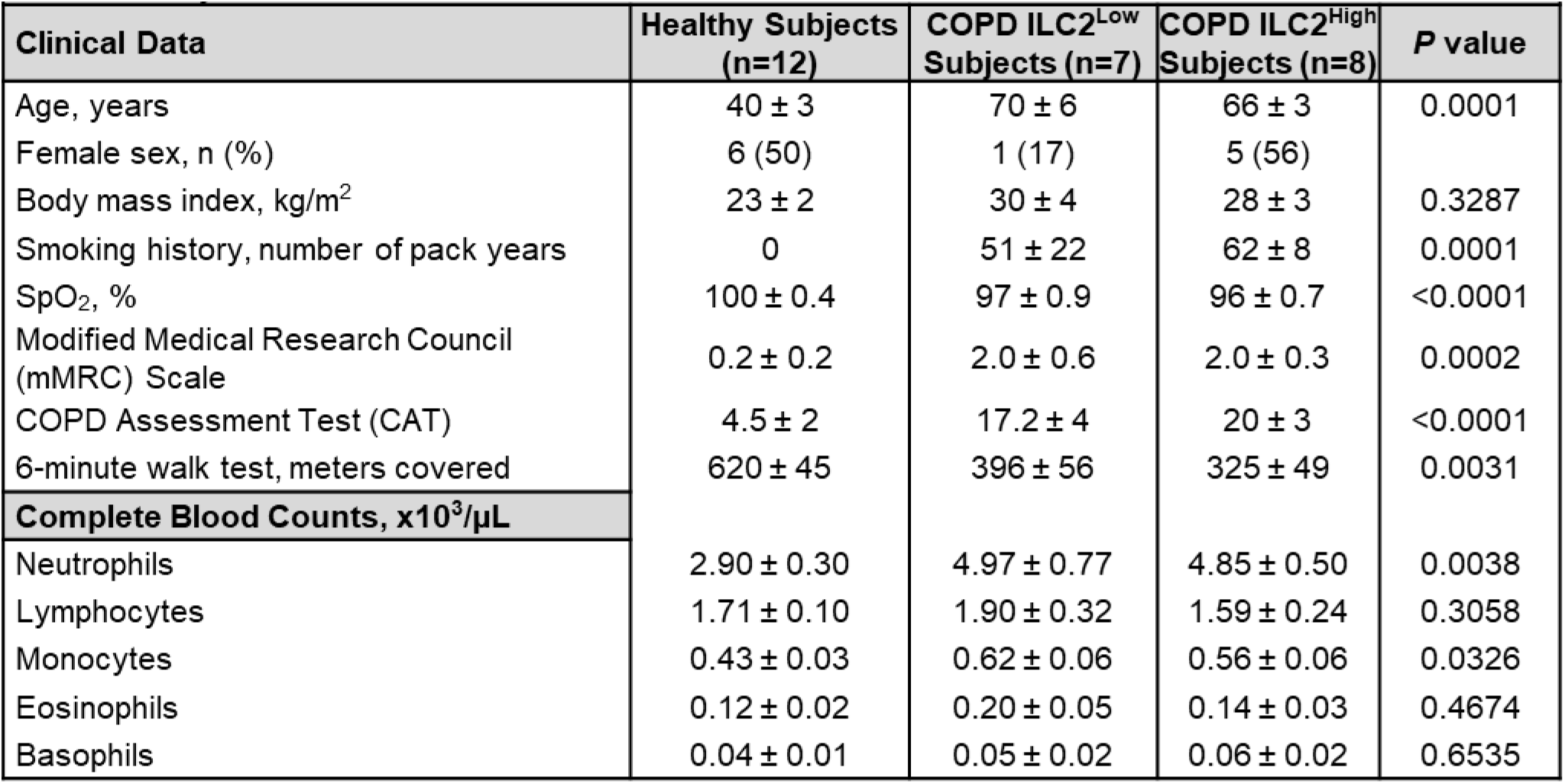
Characteristics of healthy subjects and patients with ILC2^low^ and ILC2^high^ COPD sputum. Clinical data and complete blood counts (cells x10^3^/µL) of n=12 healthy subjects and COPD patients with ILC2^low^ (n=7) and ILC2^high^ (n=8; defined as more than 1500 ILC2/mL) sputum. Data are presented as mean ± SEM. *P* values calculated as ANOVA for continuous normally distributed values.

| **Table 1. Characteristics and Peripheral Blood Cell Counts of Healthy and COPD ILC2<sup>Low</sup> and COPD ILC2<sup>High</sup> Subjects** |  |  |  |  |
| --- | --- | --- | --- | --- |
| **Clinical Data** | **Healthy Subjects (n=12)** | **COPD ILC2<sup>Low</sup> Subjects (n=7)** | **COPD ILC2<sup>High</sup> Subjects (n=8)** | **P value** |
| Age, years | 40 ± 3 | 70 ± 6 | 66 ± 3 | 0.0001 |
| Female sex, n (%) | 6 (50) | 1 (17) | 5 (56) |  |
| Body mass index, kg/m<sup>2</sup> | 23 ± 2 | 30 ± 4 | 28 ± 3 | 0.3287 |
| Smoking history, number of pack years | 0 | 51 ± 22 | 62 ± 8 | 0.0001 |
| SpO<sub>2</sub>, % | 100 ± 0.4 | 97 ± 0.9 | 96 ± 0.7 | <0.0001 |
| Modified Medical Research Council (mMRC) Scale | 0.2 ± 0.2 | 2.0 ± 0.6 | 2.0 ± 0.3 | 0.0002 |
| COPD Assessment Test (CAT) | 4.5 ± 2 | 17.2 ± 4 | 20 ± 3 | <0.0001 |
| 6-minute walk test, meters covered | 620 ± 45 | 396 ± 56 | 325 ± 49 | 0.0031 |
| **Complete Blood Counts, x10<sup>3</sup>/μL** |  |  |  |  |
| Neutrophils | 2.90 ± 0.30 | 4.97 ± 0.77 | 4.85 ± 0.50 | 0.0038 |
| Lymphocytes | 1.71 ± 0.10 | 1.90 ± 0.32 | 1.59 ± 0.24 | 0.3058 |
| Monocytes | 0.43 ± 0.03 | 0.62 ± 0.06 | 0.56 ± 0.06 | 0.0326 |
| Eosinophils | 0.12 ± 0.02 | 0.20 ± 0.05 | 0.14 ± 0.03 | 0.4674 |
| Basophils | 0.04 ± 0.01 | 0.05 ± 0.02 | 0.06 ± 0.02 | 0.6535 |

To understand if age was playing a significant role in the differences observed between healthy, ILC2^low^, and ILC2^high^ subjects, we performed regression correcting for age (**Table 2**). FEV_1_ and sputum neutrophils and eosinophils, correlated with sputum ILC2s after correcting for age. CAT score showed a correlation that failed to reach statistical significance when corrected for age (p=0.051), while FVC failed to reach significance before and after the adjustment. To understand the mechanistic relationship between ILC2s, SP-D, and IL-17A, we used mouse and *ex vivo* models.

**Table 2.** Regression models for clinical characteristics in COPD. FEV_1_, (% predicted), FVC (% predicted), and CAT score were analyzed by linear regression. Sputum neutrophils and eosinophils were analyzed by logistic regression. Data are presented as a β coefficient or odds ratio, with 95% confidence interval and p value.

| Table 2. Regression models for clinical characteristics in COPD |  |  |  |
| --- | --- | --- | --- |
| Outcome variable | Predictor variable | Unadjusted model | Adjusted (age) model |
| FEV<sub>1</sub>, % predicted | Sputum ILC2s | -21.7 (7.02, -3.09) p=0.0053 | -17.4 (6.51, -2.67) p=0.014 |
| FVC, % predicted | Sputum ILC2s | -10.7 (5.92, -1.81) p=0.083 | -8.63 (6.03, -1.43) p=0.17 |
| CAT score | Sputum ILC2s | 7.09 (2.82, 2.52) p=0.019 | 5.63 (2.72, 2.07) p=0.051 |
| Sputum neutrophils | Sputum ILC2s | 1.37 (0.38, 3.61) p=0.0015 | 1.25 (0.39, 3.21) p=0.0042 |
| Sputum eosinophils | Sputum ILC2s | 10.46 (3.62, 2.88) p=0.0086 | 8.45 (3.46, 2.44) p=0.023 |

### ILC2 counts in ILC2^high^ (but not ILC2^low^) sputum samples are correlated with SP-D release to sputum and serum SP-D leakage

The relationship between SP-D and ILCs has not been fully characterized. Given the previous observations that airway SP-D is diminished in subjects with COPD and SP-D^-/-^ mice additionally develop spontaneous airway inflammation and activation of pulmonary innate immune cells, we sought to clarify the role of ILCs in COPD disease severity and understand how SP-D impacts disease pathogenesis and ILC activation.

Sputum SP-D measurements were elevated in ILC2^high^ compared to ILC2^low^, and both ILC2^high^ and ILC2^low^ COPD subjects had significantly higher concentrations of sputum SP-D compared to healthy controls (**Fig. 3A**). Since prior studies demonstrated that lack of SP-D in humans was associated with COPD (16) and in mice was associated with the spontaneous development of COPD (18, 21), we hypothesized that elevated sputum SP-D in ILC2^high^ subjects was nonfunctional. Indeed, prior studies have shown that the tertiary structure of SP-D is subject to post-translational modifications which leads to unraveling of the dodecamer, rending its immunosuppressive capability nonfunctional (22, 23). Here we found that while overall SP-D levels were increased in ILC2^high^ subjects, there was significant degradation of the native SP-D structure, suggesting that it was nonfunctional in these subjects (**Fig. 3B**). To study the relationship of COPD severity and elevated ILC2s and SP-D, we correlated sputum ILC2 counts with SP-D concentrations and found that the most highly correlated was the ILC2^high^ group (**Fig. 3C**). Positive linear correlations of total sputum SP-D with sputum CCL24 and IL-5 were also found with the ILC2^high^ group (**Fig. 3D**).

**Figure 3.**
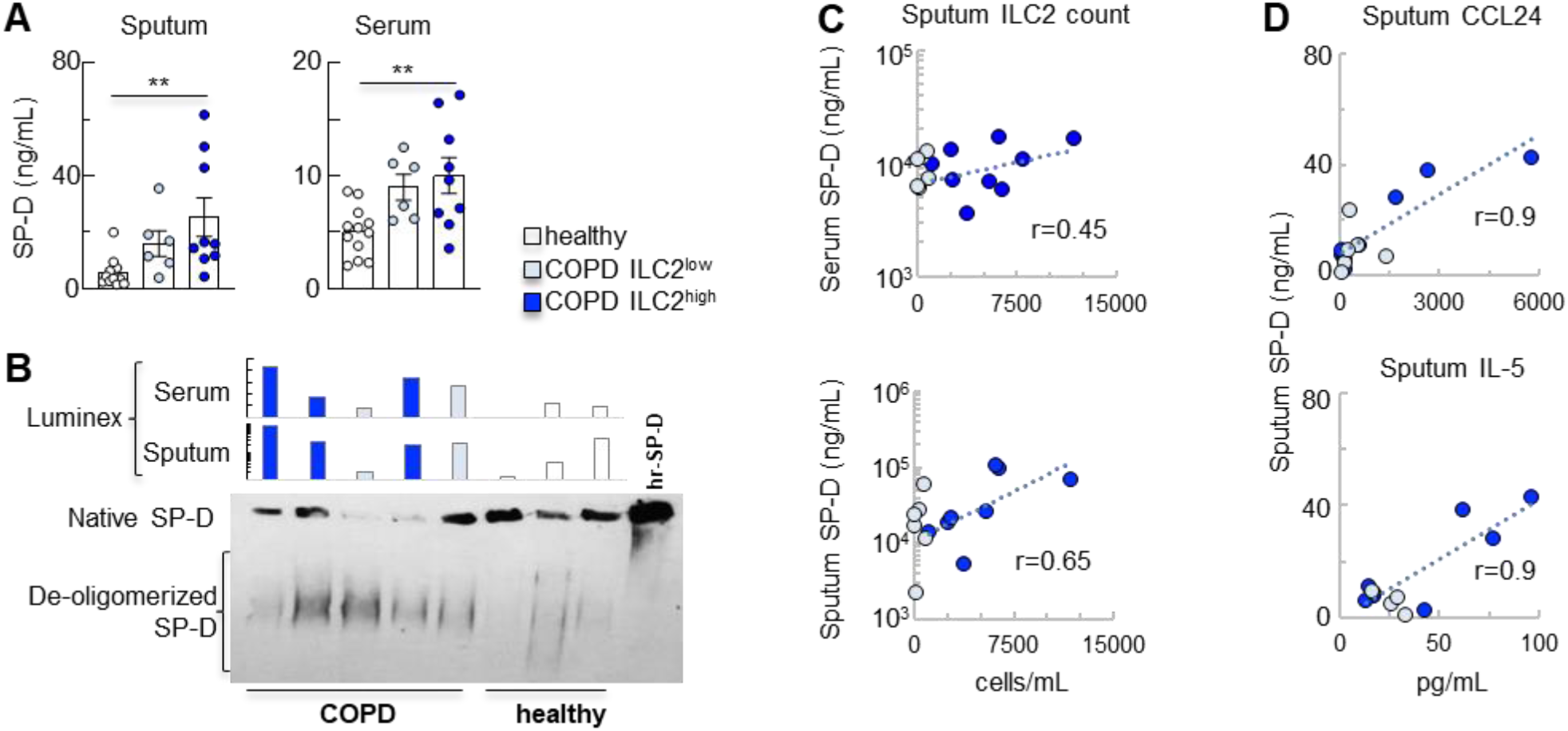
ILC2 counts in ILC2^high^ (but not ILC2^low^) sputum samples are correlated with SP-D release to sputum and serum SP-D leakage. **(A)** SP-D measured by Luminex^®^ Assay in the cell free supernatant of sputum (left panel) and serum samples from healthy subjects and COPD patients with ILC2^low^ and ILC2^high^ sputum (ng/mL). Mean±SEM of n=12 healthy, and ILC2^low^ and ILC2^high^ samples from n=7 and n=8 COPD subjects, respectively. **p=0.003 Sputum; **p=0.007 Serum; Kruskal Wallis multiple comparison. **(B)** SP-D Native gel electrophoresis (Representative sputum samples, bottom panel) using an in-house developed mouse anti-SPD (1µg; 106FE12 monoclonal), followed by goat anti-mouse conjugated HRP 1:3000. De-oligomerized low molecular weight components of SP-D streak down the native gel, while the intact (native) SP-D molecule stays on the top. Corresponding Luminex^®^ sputum and serum samples denote the extent of serum leakage of SP-D from the same individuals. **(C)** Correlations of total serum (top) and sputum (bottom) SP-D and sputum ILC2 counts in ILC2^high^ (dark blue circles) sputum samples from COPD subjects. **(D)** Correlations of total sputum SP-D (Luminex^®^) and sputum CCL24 (Eotaxin 2, top panel) and IL-5. **(C-D):** Linear regression (r: Pearson’s correlation coefficient); only ILC2^high^ samples included. There was no correlation observed in the ILC2^low^ samples.

Interestingly, there was no difference in cumulative number of pack years smoked between ILC2^high^ and ILC2^low^ individuals, suggesting that overall smoke exposure wasn’t playing a strong role in the clinical presentation of ILC2^high^ subjects (**Fig. E1**).

### Effect of SP-D deficiency on airway inflammation and lung ILC2s

While SP-D was previously suggested to play an important role in COPD pathogenesis and capable of inhibiting immune cells (10, 12), its effects on ILC2s have never been evaluated. Mice with genetically ablated SP-D expression (SP-D^-/-^) spontaneously develop emphysema, a cardinal feature of COPD (18). Because the neutrophil plays a prominent role in the pathogenesis of COPD (24), we hypothesized that SP-D^-/-^ mice would also spontaneously develop airway neutrophilia. At 6 weeks of age, SP-D^-/-^ mice showed marked BAL neutrophilia that was completely absent in C57BL/6 wild-type (WT) mice (**Fig. 4A**). This was accompanied by significantly increased BAL KC, the mouse equivalent of IL-8 (**Fig. 4B**). Given that the native SP-D structure was not maintained in the sputum of our COPD subjects, these SP-D^-/-^ data faithfully recapitulate our clinical observations. We therefore studied the presence and function of lung ILC2s in the absence of SP-D.

**Figure 4.**
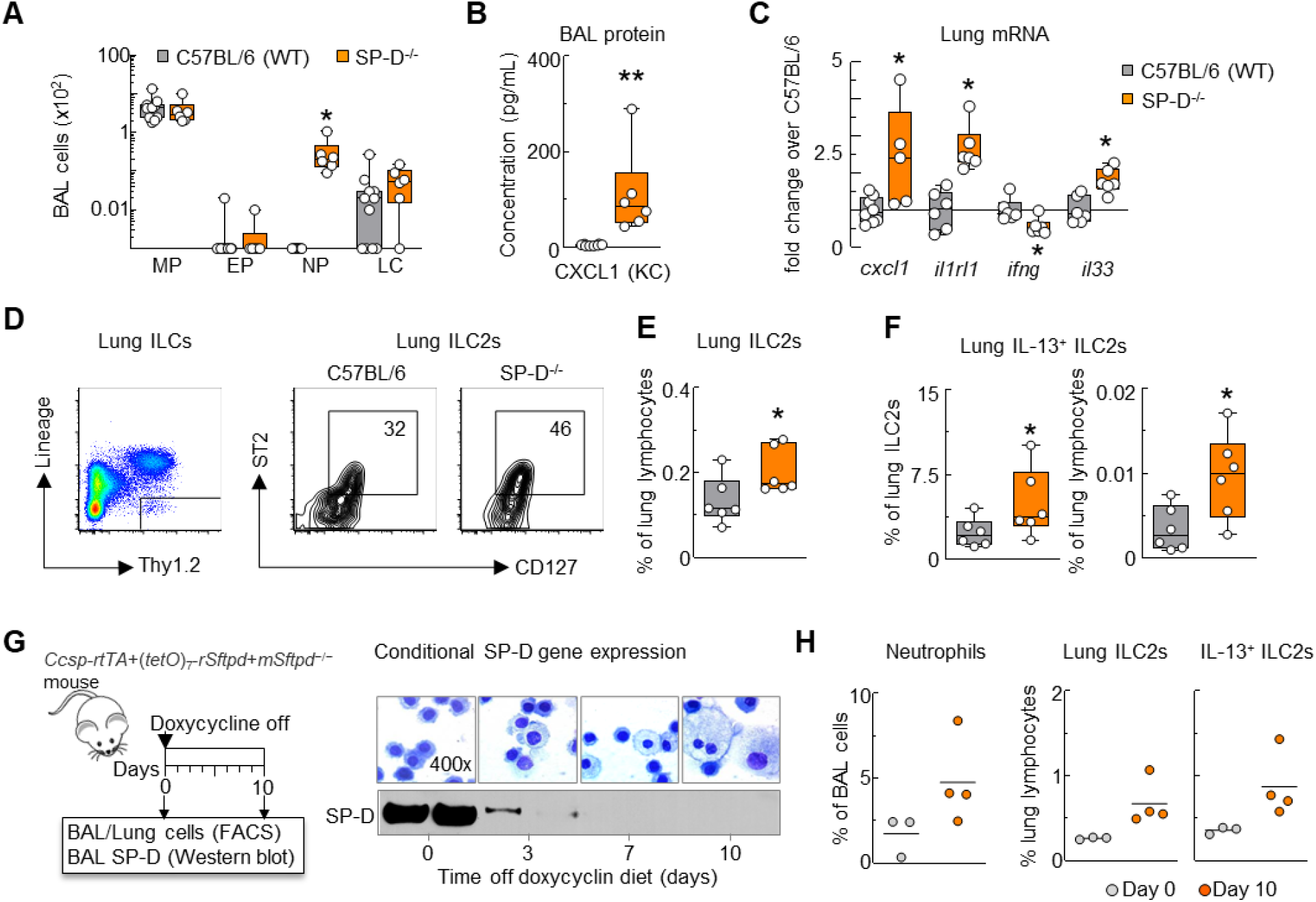
Effect of SP-D deficiency on airway inflammation and lung ILC2s. **(A)** 8-12 weeks old age and sex matched SP-D-/- and wild type (C57BL/6) littermates were studied for BAL and lung cells and cytokines. Cytospins of BAL cells were stained with Kwik-Diff and assessed for the presence of macrophages (MP), eosinophils (EP), neutrophils (NP) and lymphocytes (LC). Differential BAL cell counts (x10^2^) were calculated using the total BAL cells. **(B)** Cell-free BAL supernatant was assessed by ELISA for KC concentration (pg/mL). **(C)** Total lung RNA was extracted and processed for cxcl1, il1rl1, ifng and il33 mRNA expression by qPCR. (fold change over C57BL/6 average ctrl values). **(D)** Gating strategy for mouse lung ILCs: live Lin^-^Thy1.2^+^ cells (lineage markers – CD3, CD4 CD5, CD11c, GR-1, B220, DX5). Representative scatter plots showing the proportion of lung ILCs in that were CD127^+^ST2^+^ (ILC2s). **(E)** Quantification of lung ILC2s (% of lung lymphocytes). **(F):** Intracellular IL-13 expression by lung ILC2s. Mean±SEM of n=6-11 **(A)**, n=6-8 **(B, C, E)**; *p<0.05 or **p<0.01, Student’s *t*-test (C57Bl/6 vs. SP-D^-/-^). **(G):** In conditional SP-D expressing (*Ccsp-rtTA*+(*tetO*)_7_-*rSftpd*+*mSftpd*^−/−^) mice, expression of the SP-D gene was tied to a tetracycline promoter in club cells of the lung. Doxycycline diet was removed on day 0; mice were studied on days 0 and 10. SP-D was detected in the BAL supernatant (western blot) and cytospins of BAL cells from corresponding samples were assessed (Kwik-Diff, 400x). **(H):** Elimination of SP-D expression led to airway neutrophilia and activation of lung ILC2s (FACS analysis). Quantification of lung ILC2s and IL-13^+^ ILC2s (% of lung lymphocytes) n=3-4.

Compared to WT mice, SP-D^-/-^ displayed significantly elevated mRNA expression of cxcl1, il1rl1 and *Il33* in the lung (**Fig. 4C**). We thus hypothesized that ILC2s, a target of IL-33 activation (2), would be elevated in the SP-D^-/-^ lung. Innate lymphoid cells (ILCs) derived from whole lung homogenate were Lineage^-^Thy1.2^+^, furthered characterized as ILC2s by expression of CD127 and ST2 (the IL-33 receptor) (**Fig. 4D**). SP-D^-/-^ mice had significantly more lung ILC2s than WT mice, suggesting that ILC2s were a direct target of SP-D immunomodulation (**Fig. 4E**). Intracellular IL13 was measured and more highly expressed in ILC2s from SP-D^-/-^ mice (**Fig. 4F**).

In conditional SP-D expressing (*Ccsp-rtTA*+(*tetO*)_7_-*rSftpd*+*mSftpd*^−/−^) mice, expression of the SP-D gene was tied to a tetracycline promoter in club cells of the lung. Doxycycline diet was removed on day 0; mice were studied on days 0 and 10. SP-D was detected in the BAL supernatant (western blot) and cytospins of BAL cells from corresponding samples were assessed (**Fig. 4G**). Elimination of SP-D expression led to airway neutrophilia and activation of lung ILC2s and intracellular IL13 expression as quanitifed by FACS analysis (**Fig. 4H**).

### SP-D suppressed the emergence of IL-13 and IL-17A-producing ILC2s in the lung

Given that our ILC2^high^ COPD subjects displayed significant sputum neutrophilia and elevated IL-17A+ ILCs, and SP-D^-/-^ mice had elevated airway neutrophils, we hypothesized that SP-D regulated IL-17A in ILC2s. We FACS-sorted Lineage^-^ Thy1.2^+^CD25^+^ST2^+^ cells (**Fig. 5A**). Purity of ILC2 sorts were confirmed by flow cytometry immediately following ILC2 isolation (**Fig. E2** in the online data supplement). Analysis of gata 3 and IL-13 expression demonstrated significantly increased expression of IL-13 in SP-D^-/-^ mice and rSP-D dose-dependently inhibited IL-13 expression (**Fig. 5B, C and D**). ILC2s were also sorted for RORγt expression and shown to be increased significantly in SP-D^-/-^ mice (**Fig. 5E, F,** G and H). Strikingly, SP-D^-/-^ ILC2s secreted IL-17A that was completely absent from WT ILC2s and was dose-dependently inhibited by rSP-D *ex vivo* (**Fig. 5I and J**). These results indicate that SP-D controls IL-17A expression in lung ILC2s.

**Figure 5.**
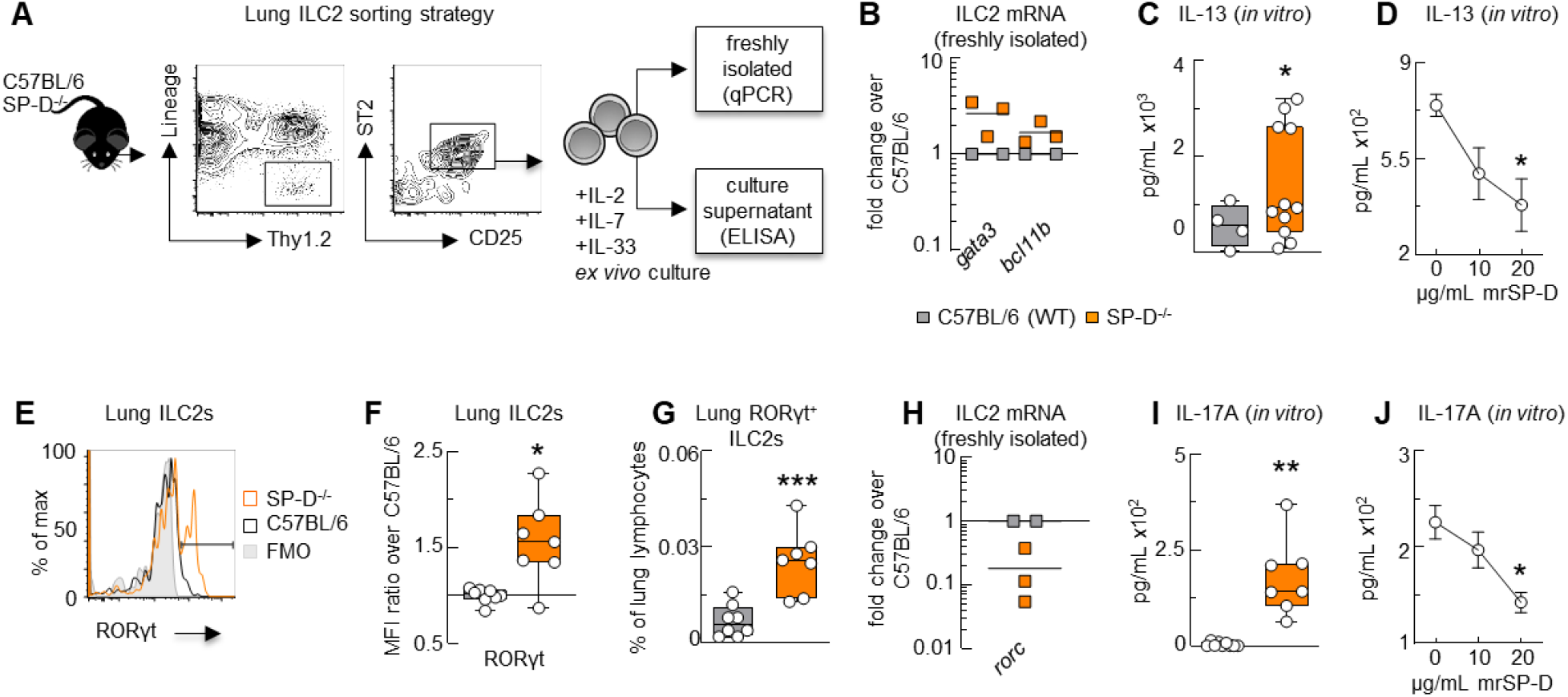
SP-D suppressed IL-13 and IL-17A co-expressing inflammatory ILC2s. **(A):** ILC2s (live CD45^+^Lineage^-^Thy1.2^+^ST2^+^CD25^+^ cells) were freshly isolated from mouse lung for mRNA analysis, or *in vitro* culture in 10 ng/mL IL-2+7+33. **(B, H):** Gene expression (qPCR) analysis of freshly isolated ILC2s (log fold change over C57BL/6 wild type control). **(C, I):** IL-13 and IL-17A of ILC2 supernatant after 7-day culture with 10 ng/mL IL-2+7+33 (ELISA; pg/mL). **(D, J):** IL-13 and IL-17A of ILC2 culture supernatant after addition of mrSP-D for 3 days (ELISA; pg/mL). **(E)** Overlay of RORγt expression (MFI) on lung ILC2s. **(F)** RORγt MFI in lung ILC2s (ratio over C57BL/6). **(G):** Quantification of lung RORγt^+^ ILC2s (% of lung lymphocytes). Mean **(B, H)**, quartiles (boxes) and range (whiskers) **(C, F, G, I)**, or mean±SEM **(D, J)** of n=2-3 from 3-4 pooled mice per point, n=4-11 **(C)**, n=7-9 **(D, I, J)**, n=7-8 **(E-G)**, n=7-9 **(I)**; *p<0.05, **p<0.01, ***p<0.001 (**C, F, G, I:** Student’s *t*-test; **D, J:** One-way ANOVA with Bonferroni correction).

### SP-D deficiency predisposed mice to O_3_-induced airway neutrophilia and lung ILC2 activation

The progressive nature of COPD is frequently aggravated by exacerbations. These short bouts of severe symptoms are driven, in part, by the presence of highly activated neutrophils in the airways (24, 25). Although the major cause of exacerbations are respiratory viruses that trigger airway neutrophilia (26), it is thought that COPD patients are uniquely susceptible to neutrophilic exacerbation driven by the air pollutant ozone (O_3_) (27, 28). We hypothesized that a trigger of COPD exacerbation would enhance the IL-17A signal in lung ILC2s of SP-D^-/-^ mice. WT and SP-D^-/-^ mice were exposed to 3 ppm O_3_ for 2 hours, then studied 12 hours later. SP-D^-/-^ mice showed significantly heightened airway neutrophilia after O_3_ exposure compared to WT mice (**Fig. 6A-B**). Likewise, BAL KC was induced by O_3_ exposure, elevated in the absence of SP-D (**Fig. 6C**). We next examined the impact of O_3_ on lung ILC2s. We found that SP-D^-/-^ ILC2s increased in the lung following O_3_ exposure (**Fig. 6D**). In contrast to our examination of ILC2s in SP-D^-/-^ lungs, here we used GATA3 in our gating strategy. This was required because O_3_ exposure reduced the expression of ST2 on lung ILC2s in both strains (**Fig. 6E**). This was accompanied by an O_3-_induced significant increase in ST2^low^RORγt^+^ ILC2s in the lung of SP-D^-/-^ mice (**Fig. 6F**). These data suggested that a trigger of COPD exacerbation could enhance the IL-17A signal in lung ILC2s.

**Figure 6.**
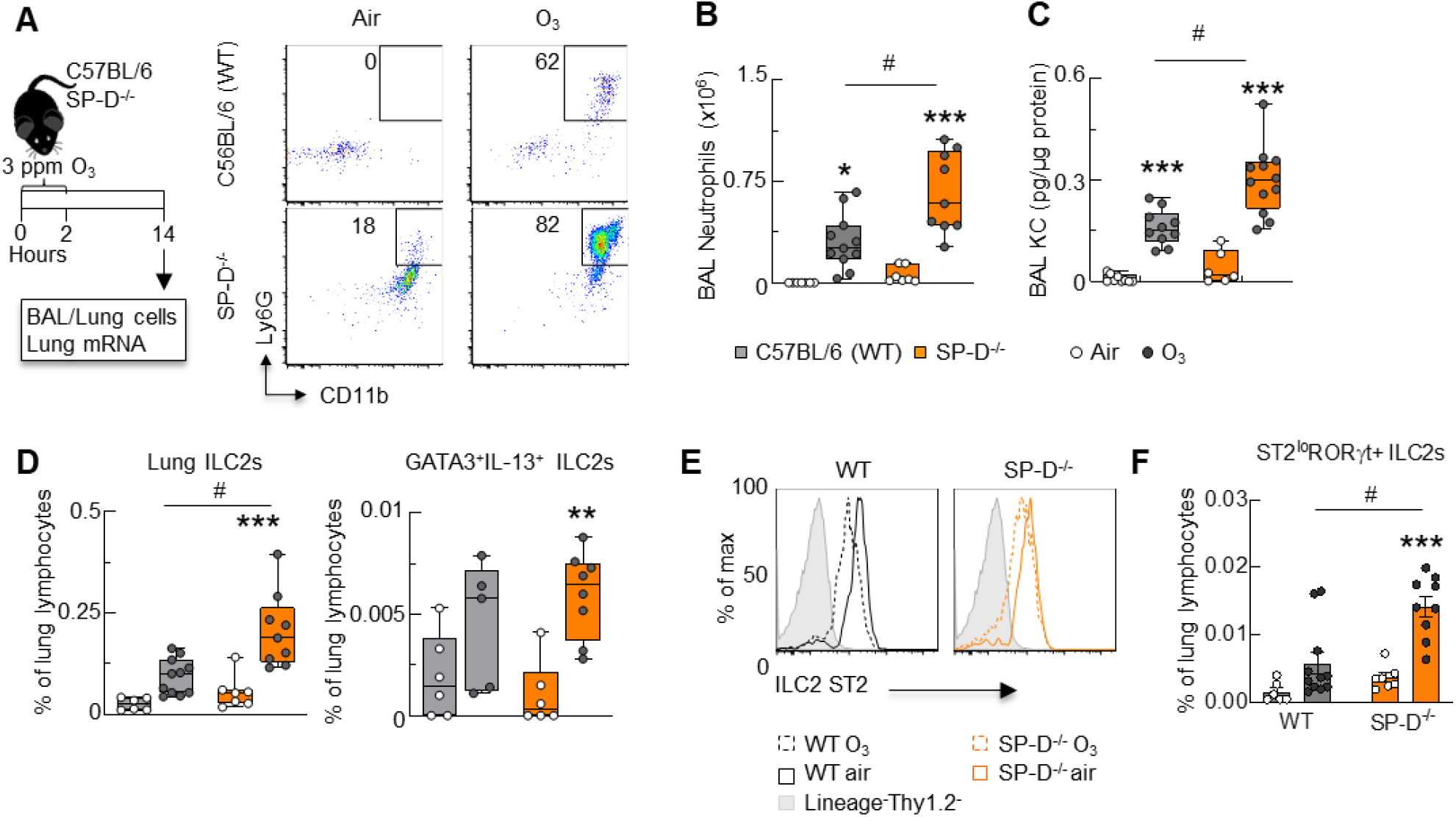
SP-D deficiency enhanced O_3_-induced airway neutrophilia and lung ILC2 activation. **(A)** Mice were exposed to 3 ppm O_3_ for 2 hours, then studied 12 hours later. Representative scatter plots depicting the proportion of live CD45^+^ BAL cells that were CD11b^+^Ly6G^+^. **(B)** Quantification of BAL neutrophils, absolute count. **(C)** KC in cell-free BAL supernatant was measured by ELISA, concentration (pg/mL). **(D)** Quantification of lung ILC2s (live Lineage^-^Thy1.2^+^CD127^+^GATA3^+^ cells), % of lung cells. **(E)** Representative overlays of ST2 expression on lung ILC2s, % of max. **(F)** Quantification of lung ST2^lo^RORγt^+^ ILC2s, % of lung cells. Mean±SEM of n=5-7 **(B)**, n=6-12 **(C)**, n=6-11 **(D)**; *p<0.05, **p<0.01, ***p<0.001, #p<0.05, Two-way ANOVA with Bonferroni’s multiple comparisons test (asterisk indicates air vs O_3_; hashmark indicates WT vs SP-D^-/-^).

### ILC2 derived IL-17A was essential for ozone-induced neutrophilia in mice

Because the COPD exacerbation trigger O_3_ caused neutrophilic airway inflammation and induced markers of IL-17A expression in lung ILC2s, we investigated if the IL-17A capability of SP-D^-/-^ ILC2s was enhanced in this model. In SP-D^-/-^ mice, O_3_ increased the proportion of lung ILC2s that expressed IL-17A compared to WT controls (**Fig. 7A**). This was accompanied by an O_3_-induced increase in the number of IL-17A^+^GATA3^+^RORγt^+^ ILC2s in SP-D^-/-^, but not WT mice (**Fig. 7B**). To understand if ILC2-derived IL-17A was important to airway neutrophilia, we employed Rag2/γc^-/-^ mice (on the Balb/c background) lacking innate and adaptive lymphocytes. Compared to Balb/c mice, Rag2/γc^-/-^ mice exhibited reduced airway neutrophilia, which could be restored with adoptive transfer of ILC2s expanded *ex vivo* prior to O_3_ (**Fig. 7C**). Correlation between % lung ILC2s and BAL neutrophils in the Rag2/γc^-/-^ +O_3_+ILC2 experimental group. R: Pearson’s correlation coefficient (**Fig 7D**). Recipient Rag2/γc^-/-^ mice given ILC2s and injected with αIL-17A had less airway neutrophilia (**Fig 7E**), expressed lower CXCL-1 and higher IL-27 compared to animals receiving αIgG1 control (**Fig 7F**). These data suggested that SP-D plays a key role in regulating the development of IL-17A^+^ and highly proinflammatory dual-positive ILC2s in the lung, and further that this process is required for airway neutrophilia.

**Figure 7.**
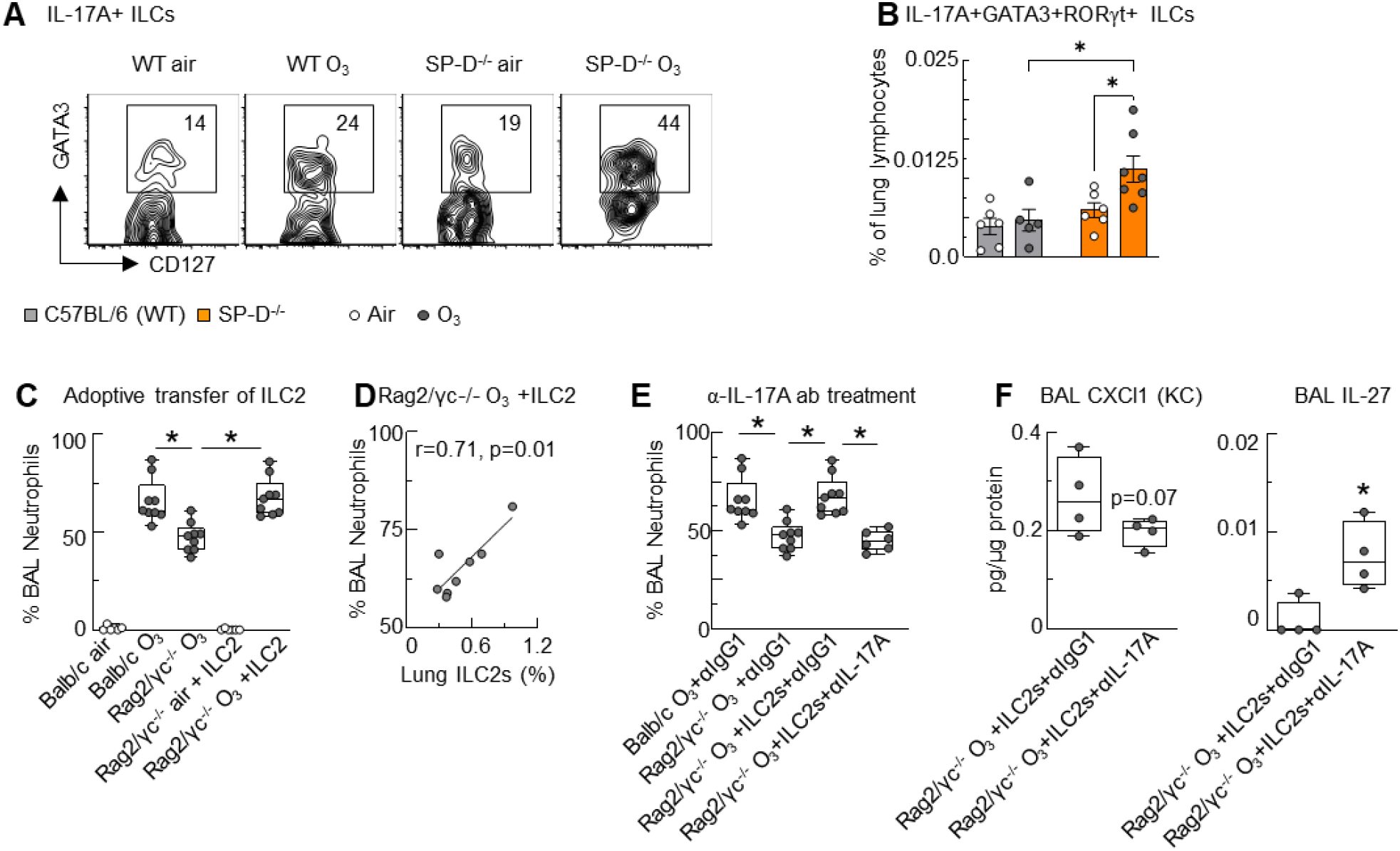
ILC2 derived IL-17A was essential for ozone-induced neutrophilia in mice. **(A-B)** SP-D deficiency heightened O_3_-induced IL-17A production by ILC2. (A): Representative scatter plots of lung IL-17A^+^ ILCs (live Lineage^-^IL-17A^+^CD127^+^Thy1.2^+^ cells) that were GATA3^+^. **(B)** Quantification of lung IL-17A^+^GATA3^+^RORγt^+^ ILC2s, % of lung cells. **(C)** Adoptive transfer of ILC2s restored O_3_-induced BAL neutrophilia: FACS sorted ILC2s from lungs of donor mice were expanded for 14 days *ex vivo* before adoptive transfer of 3x10^5^ cells into recipient Rag2/γc^-/-^ mice followed by O_3_ exposure. Quantification of BAL total cells and neutrophils by differential cell count of Diff-Quik stained cytospins. *p<0.05, air *vs*. O_3_ with or without adoptive transfer of ILC2s; #p<0.05 Balb/c *vs.* Rag2/γc^-/-^ mice exposed to O_3_ (ANOVA with Tukey’s post hoc test). **(D)** Correlation between % lung ILC2s and BAL neutrophils in the Rag2/γc^-/-^ +O3+ILC2 experimental group. r: Pearson’s correlation coefficient. **(E-F)** Mice were treated with 250 µg αIgG1 or αIL-17A *i.p.* 8, 32, and 56 hours before O3 exposure. **(E)** % BAL neutrophils counted on Kwik-Diff stained cytospins from the indicated groups. **(F)** BAL CXCL1 and IL-27 were measured by ELISA. Data expressed as mean±SEM of n=5-9 and are representative of at least two independent experiments. *p<0.05 (ANOVA with Tukey’s post hoc test).

**Figure 8.**
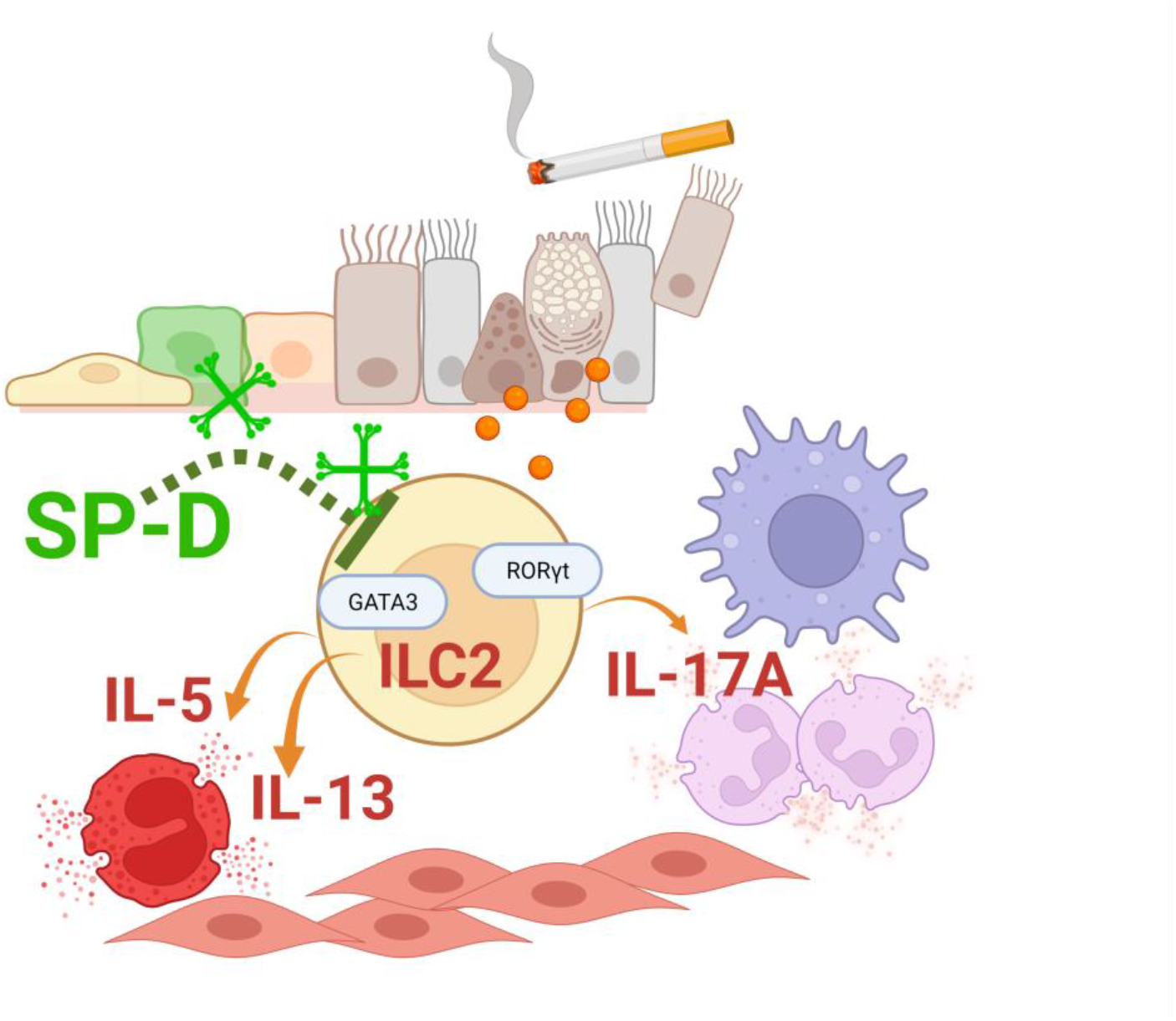
IL-17A production in lung ILC2s is driven by environmental exposures and is counteracted by SP-D. We propose that SP-D suppresses activation and plasticity of GATA3+RORγt+ IL-13/IL-17A producing lung ILC2s, thereby protecting from eosinophilic and neutrophilic airway inflammation in COPD.

## DISCUSSION

Here we establish a mechanistic link between SP-D and lung ILC2s, complementing our clinical studies wherein subjects with elevated sputum ILC2s were severely ill and had degraded SP-D. ILC2^high^ subjects with COPD had decreased FEV_1_, increased CAT scores, and enhanced sputum neutrophilia and eosinophilia compared to ILC2^low^ and healthy subjects. This was accompanied by the emergence of ILCs co-expressing IL-5 and IL-13 with IL-17A. Mechanistically, lack of SP-D expression in the mouse lung led to IL-17A production in lung ILC2s that was associated with airway neutrophilia. SP-D^-/-^ mice showed heightened susceptibility to O_3_-induced neutrophilia, which coincided with increased numbers of IL-17A^+^ lung ILC2s. These data demonstrate that sputum ILC2s may be a good predictor of disease severity in COPD, especially when mixed neutrophilic and eosinophilic inflammation is observed.

Initially, we set out to define the role of ILCs in the disease severity of COPD. Surprisingly, we found that ILCs expressing GATA3 were increased in the sputum of subjects with COPD compared to healthy controls. Within the COPD group however, we noticed that some patients clearly showed elevated ILC2s/mL sputum, while others had similar levels when compared to healthy controls. Thus, to gain deeper insight into the relevance of ILC2s in COPD, we stratified subjects into ILC2^high^ and ILC2^low^ subgroups based on the upper limit of the 95% confidence interval in healthy controls (1427 cells/mL sputum). The ILC2^high^ group had more severe disease than ILC2^low^ subjects with COPD based on lung function, CAT score, and sputum inflammation. Since ILC2s were previously recognized for their role in Type 2 immunity (2, 5), it was unexpected that they were elevated in the sputum of COPD subjects showing an IL-8/neutrophil-high clinical presentation. However, these ILC2^high^ patients displayed elevated sputum neutrophils and eosinophils, which wasn’t statistically apparent when viewing the COPD group as a whole. ILCs expressing IL-5 and IL-13 in combination with IL-17A in these subjects suggested that ILC2s gained a dual-functional phenotype, contributing to the mixed neutrophilic and eosinophilic inflammatory profile. These data suggest a novel role for ILC2s, expressing their canonical cytokines IL-5 and IL-13 in combination with IL-17A, as a highly pro-inflammatory population in severe COPD.

Prior work by Silver et al. showed that ILC1s increased and ILC2s decreased in the peripheral blood of patients with COPD, while De Grove et al. demonstrated that ILC2s were decreased and ILC3s increased in the lung tissue of COPD (7, 8). Here we studied ILCs in the sputum, and we used transcription factors to define ILCs as ILC1s, ILC2s, and ILC3s. This tissue specificity and slight modification of gating strategy could explain the discrepancies between our studies. Moreover, the observation that ILC2^high^ subjects had significant eosinophilia compared to healthy controls suggest that our cohorts were clinically different.

We wondered how ILC2s could gain IL-17A functionality. SP-D was previously recognized as an important immunomodulator in the pathogenesis of COPD (16, 18, 21, 29–31). In our clinical study, we found that while overall SP-D concentrations were increased the sputum of subjects with COPD, the native structure of the protein was unraveled. Because the SP-D structure is subject to post-translational modifications that render its immunomodulatory capabilities nonfunctional (22, 23), we propose that nonfunctional SP-D in the ILC2^high^ group led to enhanced activation of ILC2s and a mixed cytokine profile of ILCs in the sputum. To study the mechanisms of SP-D and ILC2-acquired IL-17A, we turned to mouse and *ex vivo* models.

We utilized SP-D^-/-^ mice that spontaneously develop the histological and inflammatory features of COPD (18). Lack of SP-D led to increased numbers of ILC2s and were capable of IL-17A production *ex vivo*. IL-17A secretion was dose-dependently inhibited by recombinant SP-D, suggesting that lack of functional SP-D in the sputum of COPD patients could lead to enhanced numbers of IL-17A-expressing ILC2s. Using the air pollutant O_3_ as a trigger for neutrophilic exacerbation, we found that activation of IL-17A expressing ILC2s was enhanced in the absence of SP-D. This activation was necessary for the observed airway neutrophilia, linking SP-D and ILC2s to the clinical features observed in our COPD cohort. Since the regulation of ILC2s (32, 33) is poorly understood and the vast majority of SP-D in the body is produced in the lung, these data shed light on lung-specific control of ILC2 activity. Other chronic inflammatory lung diseases characterized by impaired SP-D may predispose individuals to ILC2-mediated adverse pathologies. It is possible that therapeutic interventions to elevate functional SP-D expression may impact this pathway.

As ILC2s are among the first to respond to inhaled pathogens and environmental stressors in the lung, how they initiate the inflammatory response may have a disproportionately large implication to disease pathogenesis. In COPD, lack of functional SP-D may skew ILC2s to produce IL-17A in combination with IL-5 and IL-13, leading to a mixed inflammatory profile and more severe disease.

## Data Availability

All data produced in the present work are available upon reasonable request to the authors.

## ABBREVIATIONS

COPD: Chronic Obstructive Pulmonary Disease
SP-D: Surfactant Protein-D
ILC: Innate Lymphoid Cell
ILC1: Group 1 Innate Lymphoid Cell
ILC2: Group 2 Innate Lymphoid Cell
ILC3: Group 3 Innate Lymphoid Cell
FEV_1_: Forced Expiratory Volume in one second
FVC: Forced Vital Capacity
GOLD: Global Initiative for Chronic Obstructive Lung Disease
PFT: Pulmonary Function Testing
mMRC: Modified Medical Research Council scale
CAT: COPD Assessment Test
MP: Macrophage/monocyte
LC: Lymphocyte
NP: Neutrophil
EP: Eosinophil
WT: C57BL/6 Wild-type
BAL: Bronchoalveolar Lavage Fluid
O_3_: Ozone
PBS: Phosphate-buffed Saline
rSP-D: Mouse Recombinant SP-D
SpO_2_: Specific Oxygen

## Author Contributions

C.H.F. planned, performed, and analyzed all experiments. A.L., L.F., M.J., T.T., M.T., M.S., B.K., and A.Z. planned and performed clinical studies.

M.G.E. planned and performed mouse experiments. A.H. conceived and oversaw all aspects of the study. C.H.F., A.L., and A.H. wrote the manuscript.

## Online Data Supplement

This article has an online data supplement, which is accessible from this issues table of contents online at www.atsjournals.org.

## ACKNOWLEDGEMENTS

The authors thank members of the Haczku laboratory for their suggestions during laboratory meetings and assistance in technical aspects of the study: Erik Larson, Sarah Killingbeck, Jessica-Miranda Bustamante, and Sean Ott.

The authors thank Abigail Spinner in the Clinical Laboratory at the UC Davis California National Primate Research Center for her expertise in flow cytometry and assistance with FACS experiments. We also thank the UC Davis Air Pollution Journal Club led by Dr. Laura Van Winkle for their critical review of the manuscript.

## Online Data Supplement

### Methods

#### Human Subjects

Inclusion criteria for COPD subjects included: a life tobacco exposure of >10 pack years, incomplete reversible airflow obstruction on spirometry with a post-bronchodilator forced expiratory volume in 1 second (FEV_1_) % predicted of <80% on screening spirometry, and absence of asthma screening criteria. Disease severity was established according to Global Initiative for Chronic Obstructive Lung Disease (GOLD) classification. Subjects were excluded for age less than 18, alpha1-antitrypsin deficiency, other significant lung or major systemic illness, current use of highly active systemic immunosuppressants or high dose systemic steroidal anti-inflammatory drugs, pregnancy, or prisoner status or otherwise institutionalized. Healthy volunteers were included if they were greater than or equal to 18 years of age and excluded for any known lung disease, other significant lung or major systemic illness, pregnancy, or prisoner status or otherwise institutionalized. COPD and healthy subjects were instructed via informed consent that participation in this study was completely voluntary and that they could withdraw their participation at any time.

#### Flow cytometry antibodies, staining, and analysis

Anti-mouse antibodies purchased from BioLegend included: Trustain fcx CD16/32 (cat# 101320, clone 93), CD3-FITC (cat# 100204, clone 17A2), CD11c-FITC (cat# 117306, clone N418), CD49b-FITC (cat# 108906, clone DX5), Ly6G-FITC (cat# 108406, clone Gr-1), CD90.2-APC/Cy7 (cat# 105328, clone 30-H12), CD45-APC/Cy7 (cat# 103116, clone 30-F11), IL-17A-PE/Cy7 (cat# 506921, clone TC11-18H10.1). Anti-mouse antibodies purchased from BD Biosciences included: CD5-FITC (cat# 553021, clone 53-7.3), B220-FITC (cat# 553088, clone RA3-6B2), Ly6G-PE (cat# 551461, clone 1A8), GATA3-PECF594 (cat# 563510, clone L50-823), RORγt-BV650 (cat# 564722, clone Q31-378), CD90.2-APC (cat# 553007, clone 53-2.1), CD25-PE/Cy7 (cat# 552880, clone PC6). Anti-mouse antibodies purchased from eBioscience included: CD4-FITC (cat# 11-0041-82, clone GK1.5), ST2-PE (cat# 12-9335-82, clone RMST2-2), IL-5-PE (cat# 12-7052-81, clone TRFK5), IL-13-APC (cat# 50-7133-80, clone eBio13A), CD127-efluor450 (cat# 48-1273-82, clone eBioSB/199), CD11b-APC (cat# 17-0112-83, clone M1/70).

Anti-human antibodies purchased from BioLegend included: CD14-FITC (cat# 301804, clone M5E2), CD16-FITC (cat# 302006, clone 3G8), CD19-FITC (cat# 302206, clone HIB19), CD20-FITC (cat# 302304, clone 2H7), IL-5-PE (cat# 504304, clone TRFK5), IL-13-PerCP/Cy5.5 (cat# 501911, clone JES10-5A2), T-bet-PE/Dazzle 594 (cat# 644828, clone 4B10), and GATA3-PE (cat# 653804, clone 16E10A23). Anti-human antibodies purchased from BD Biosciences included: CD3-Alexa Fluor 488 (cat# 557705, clone SP34-2), CD56-Alexa Fluor 488 (cat# 557699, clone B159), and RORγt-BV650 (cat# 563424, clone Q21-559). Anti-human antibodies purchased from Miltenyi Biotec included: CD127-PE-Vio770 (cat# 130-113-412, clone MB15-18C9) and IL-17A-VioBlue (cat# 130-097-018, clone CZ8-23G1).

#### qPCR

The *β*-*actin* primer sequence was: Forward GCAGATGTGGATCAGCAAG, Reverse AGGGTGTAAAACGCAGCTC. The *Il33* primer sequence was: Forward GATGGGAAGAAGCTGATGGTG, Reverse TTGTGAAGGACGAAGAAGGC.

### Supplemetal Figures

**Figure E1.**
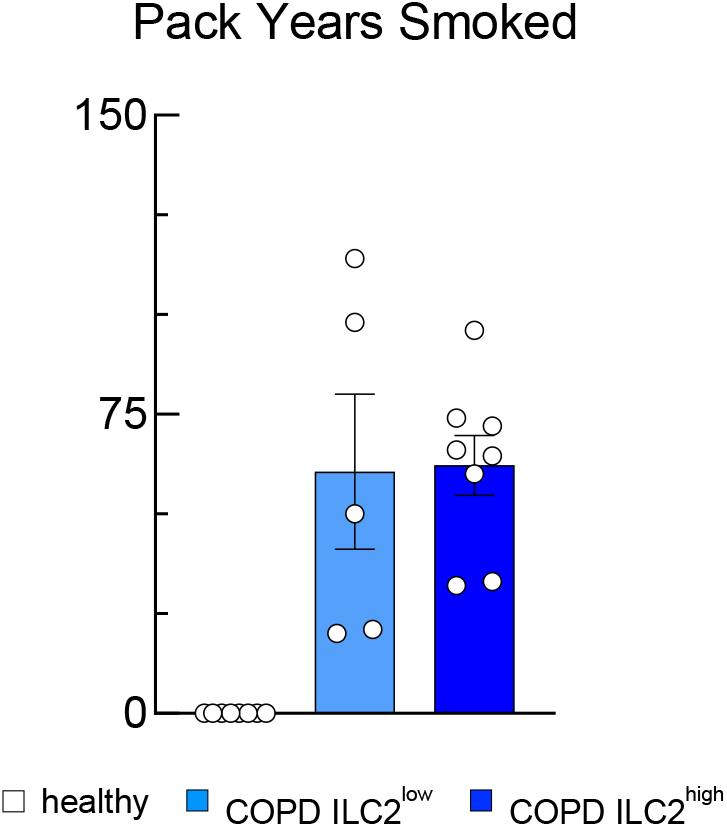
The distribution of pack years was similar between ILC2^low^ and ILC2^high^ COPD patients. The number of pack years the healthy, ILC2^low^ and ILC2^high^ COPD subjects smoked is depicted.

**Figure E2.**
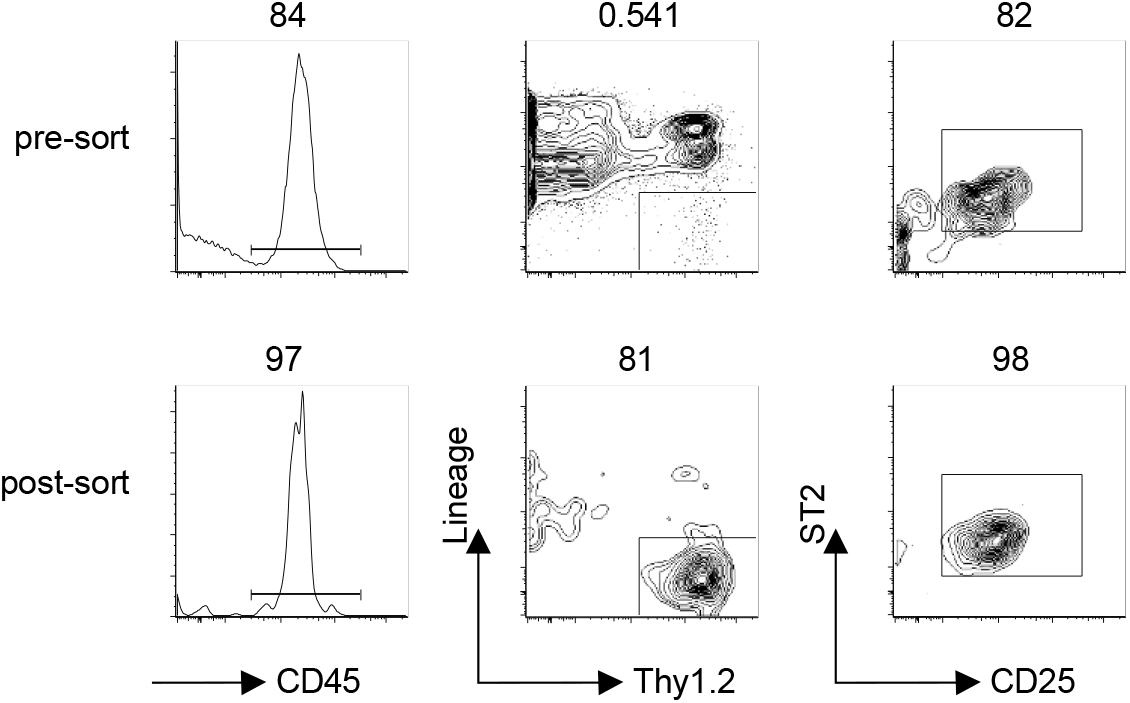
Mouse lung ILC2 sort purity. Representative scatter plots of pre-and post-sort ILC2s. Lung ILC2s were gated as live CD45^+^Lineage^-^Thy1.2^+^CD25^+^ST2^+^ cells.

